# Infection Density and Epidemic Size of COVID-19 in China outside the Hubei province

**DOI:** 10.1101/2020.04.23.20074708

**Authors:** Yukun Liu, Jing Qin, Yan Fan, Yong Zhou, Dean A. Follmann, Chiung-Yu Huang

## Abstract

The novel coronavirus (COVID-19) has spread to almost all countries in the world, claiming more than 160,000 lives and sickening more than 2,400,000 people by April 21, 2020. There has been research showing that on average, each infected person spreads the infection to more than two persons. Therefore the majority of the population is at risk of infection if no intervention measures were undertaken. The true size of the COVID-19 epidemic remains unknown, as a significant proportion of infected individuals only exhibit mild symptoms or are even asymptomatic. A timely assessment of the evolving epidemic size is crucial for resource allocation and triage decisions. In this article, we modify the back-calculation algorithm to obtain a lower bound estimate of the number of COVID-19 infected persons in China outside the Hubei province. We estimate the infection density among infected and show that the drastic control measures enforced throughout China following the lockdown of Wuhan City effectively slowed down the spread of the disease in two weeks. Our findings from China are expected to provide guidelines and enlightenment for surveillance and control activities of COVID-19 in other countries around the world.

## INTRODUCTION

In early December 2019, a cluster of pneumonia cases of unknown etiology was reported in Wuhan, a city of 11 million residents in central China. The Chinese Center for Disease Control and Prevention (China CDC) reported a novel coronavirus as the causative agent of this outbreak on January 9, 2020. To contain the spread of the virus, Wuhan, the epicenter of the coronavirus epidemic, has been placed in lockdown since January 23, 2020. The order was later expanded to the entire Hubei province in the next few days, affecting nearly 56 million people. However, it was estimated that 5 million people already left the central Chinese city, as China’s great Lunar New Year migration has already broken across the nation in the first few weeks of January. Some carried with them the new virus that has since spread throughout China and to 212 other countries, claiming almost 89,000 lives and sickening more than 1,400,000 people as of April 10, 2020. After characterizing the outbreak a Public Health Emergency of International Concern in late January, the World Health Organization (WHO) eventually declared COVID-19 as a pandemic on March 11, 2020.

Thus far, research on the basic reproduction number (R0) of COVID-19 has reported an estimated R0 in the range of 2.2 to 3.6 [1-4], which means that, on average, each infected person spreads the infection to more than two persons. Therefore, the majority of the population is at risk of infection if no intervention measures were undertaken. The true size of the COVID-19 epidemic remains unknown, as a significant proportion of infected individuals only exhibit mild symptoms or are even asymptomatic.

The intensive care needed to treat COVID-19 patients is adding pressure to the already stressed healthcare system worldwide. A recent report from WHO found that the case fatality rate was 5.8% in Wuhan, compared with 0.7% in the rest of the country [5]. The striking difference is mainly due to the sudden surge of severely ill people overwhelming the healthcare system. Hence a timely assessment of the evolving epidemic size is crucial for resource allocation and triage decisions.

In this article, we apply the back-calculation procedure developed by Brookmeyer and Gail to obtain a lower bound estimate of the number of infected individuals [6, 7]. The estimation procedure projects the observed numbers of confirmed cases to numbers previously infected, where the number of confirmed cases in each time interval follows a multinomial distribution with cell probabilities that can be expressed as a convolution of the density of the infection time and the distribution of time to diagnosis. As a result, the problem is reduced to estimating the size of a multinomial population. This approach has been applied to study infectious diseases such as acquired immunodeficiency syndrome (AIDS) and bovine spongiform encephalopathy (BSE) epidemic. (7, 8) Although the method can not predict future new infections, it provides a lower bound estimate for the number of confirmed cases in the near future to guide decision making on medical resource allocation. Moreover, the estimation of infection density provides an assessment of the infection risk during and after the drastic measure undertaken by the Chinese authorities to lockdown the city of Wuhan.

## METHODS

Let *U* denote the calendar time of COVID-19 infection for an individual and let *T* denote the incubation time from infection to diagnosis. Therefore an individual is diagnosed before a calendar time *τ* if and only if *U* + *T* ≤ *τ*. Suppose the numbers of confirmed cases of COVID-19 in a series of time intervals [*τ*_0_, *τ*_1_), [*τ*_1_, *τ*_2_), …, [*τ*_*K* − 1_, *τ*_*K*_) are available, where we set *τ*_0_ ≡−∞, so that no infection occurred prior to *τ*_0_, and *τ*_*K*_ is the date of the last available report. Assume that the distribution of *U* given *U* ≤ _*K*_ has a density function *ϕ*(*u*; ***α***), *u* ≤ τ_*K*_, where ***α*** is a *p*_*α*_-dimensional vector of parameters. Moreover, we assume that, given *U* = *u, T* is a nonnegative, continuous random variable with the distribution function *F*_*u*_(*t*; ***β***), where ***β*** is a *p*_*β*_-dimensional vector of parameters. To implement the maximum likelihood estimation, we assume that *F*_*u*_(*t*; ***β***) ≡ *F* (*t*; ***β***), that is, the time from infection to diagnosis is independent of the date when the individual was infected. This assumption may not be valid for the confirmed case in Wuhan or other cities in the Hubei province, as the medical resource in the early stage of the epidemic is extremely scarce and thus it may take a long time for infected individuals to be diagnosed. In fact, the surge in the number of confirmed cases in China on February 13 was due to a new diagnosis classification rule for cases in the Hubei province; the RT-PCR test for COVID-19 was not available for many of the previously infected individuals despite having symptoms of pneumonia. On the other hand, this assumption seems reasonable for areas outside of Hubei province as the healthcare system has not been overly stressed.

It follows from the assumption that *T* independent of *U* given *U ≤ τ*_*K*_ that the probability of being diagnosed in the time interval [*τ*_*j*−1_, *τ*_*j*_), conditioning on being infected before the last examination time *τ*_*K*_, is given by

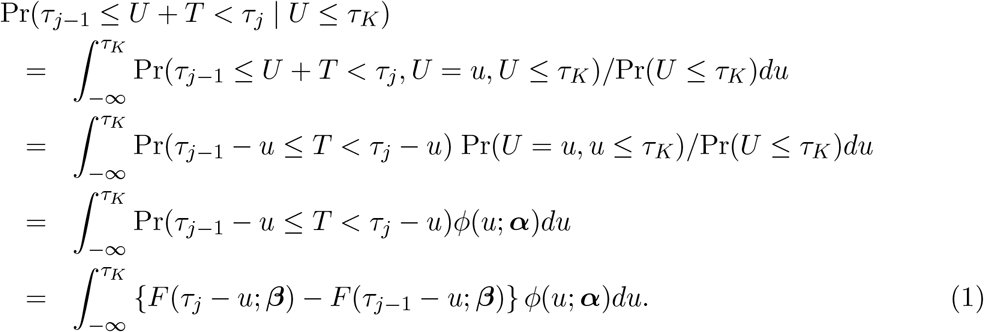

Let ***θ*** = (*α*^T^, *β*^T^)^T^ and define 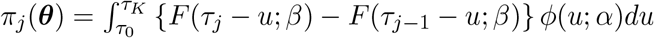.

Denote by *N* the (unknown) number of individuals infected before *τ*_*K*_ and let *X*_*j*_, *j* = 1, …, *K* be the number of cases diagnosed in the time interval [*τ*_*j−*1_, *τ*_*j*_). Define 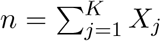, so that a total of *N − n* individuals were infected but not diagnosed before *τ*_*K*_. Define 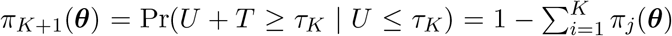. Figure 1 depicts the structure of observed data and model parameters. Write 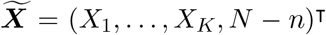 and 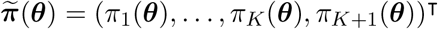, so 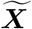 follows the multinomial distribution with trial size *N* and cell-probabilities 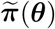. Given the observed data, *X*_1_, …, *X*_*K*_, the likelihood function of (*N*, ***θ***) is

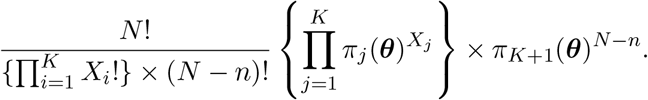

**Figure 1:**
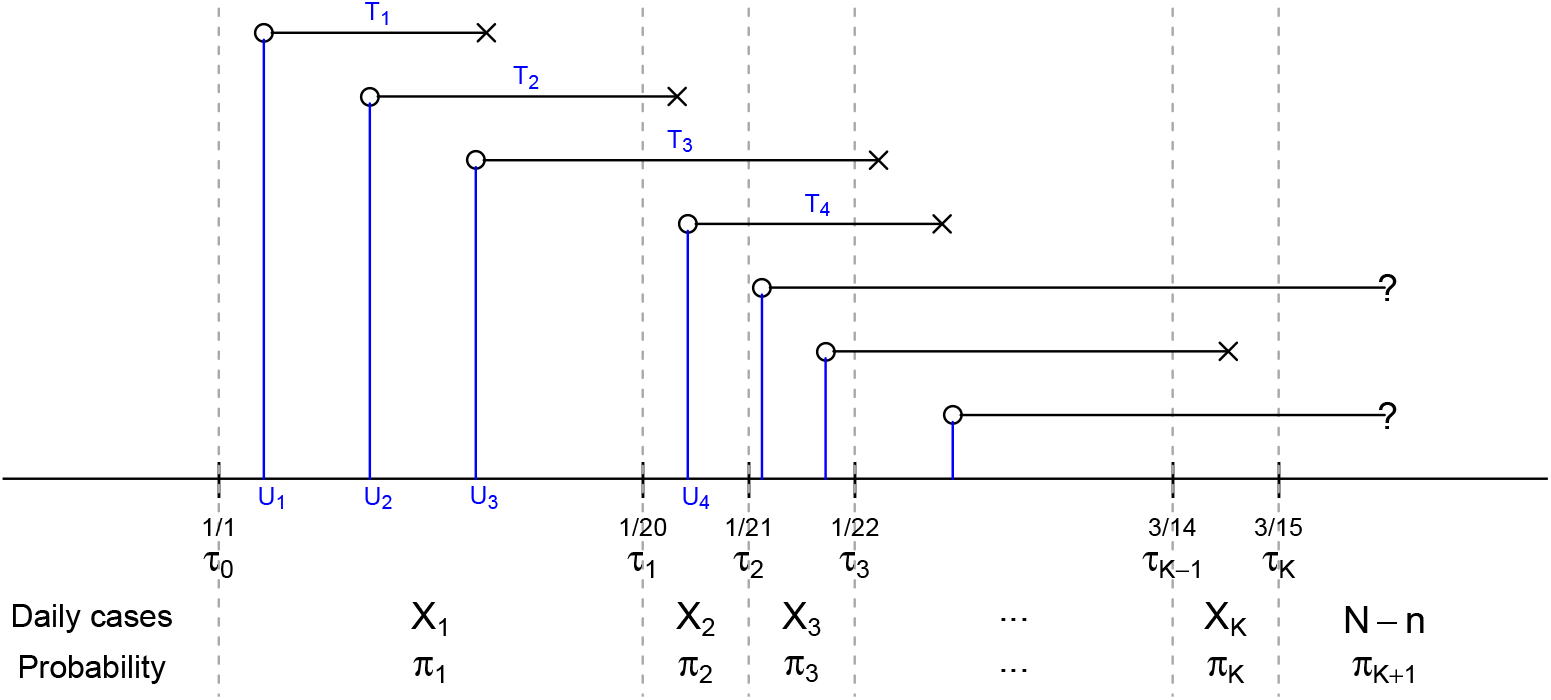
Depiction of the data structure. *U* ‘s are the calendar time of infection and *T* ‘s are the duration between infection and diagnosis. *U* ‘s and *T* ‘s are unobserved while the daily numbers of diagnosed cases *X*_1_, …, *X*_*K*_ in the *K* time intervals are observed. Here *π*_1_, …, *π*_*K*_ are the probabilities of being diagnosed in the *K* time intervals among those infected before time *τ*_*K*_.

The corresponding log-likelihood, up to a constant, is

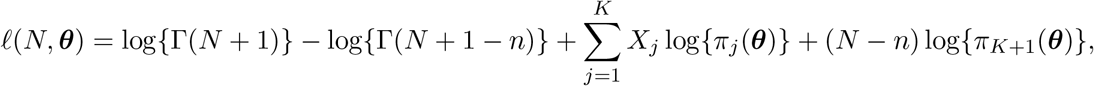

where Γ (*x*+1) = *x*! is the Gamma function. We propose to estimate (*N*, ***θ***) by the maximizer of likelihood function

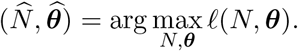

Then the expected number of confirmed cases in the time interval [*τ*_*j−*1_, *τ*_*j*_) can be estimated by 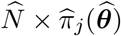, where

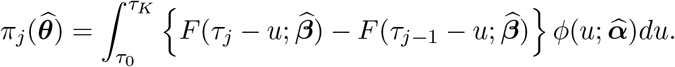

It is worthwhile to point out that the back-calculation algorithm described above estimates the two sets of parameters (***α, β***) simultaneously; this is different than the original algorithm where the parameters ***β*** in the Weibull distribution were replaced by values derived from other studies which provided information about the incubation time [6, 7]. In our analysis, *T* stands for the duration between infection and laboratory confirmation, whose information plays an important role in guiding medical resource allocation. Since no active surveillance testing and contact tracking were conducted in China before March 15, the length of time to confirmation is expected to be longer than the incubation time, that is, the duration between infection and symptom onset.

We have performed the analysis by incorporating the estimated incubation time distribution reported in [9], but the model did not fit well. As a result, we decided to estimate the two sets of parameters simultaneously. However, one major consequence of such a strategy is that the distribution of *U* and *T* are estimated subject to a location shift factor. To see this, we note that the infection time, *U*, and the time from infection to diagnosis, *T*, are not directly observed in the data, and we only get to observe *U* + *T*, that is, the time of confirmation. Hence a different set of random variables *T* ^*∗*^ = *T −δ* and *U* ^*∗*^ = *U* +*δ* yield the same distribution as *T* + *U*. On the other hand, although the location shift factor *δ* can not be estimated directly from the observed data, we can assess the magnitude of the location shift by comparing the estimated time to diagnosis distribution to what reported in the existing literature. Note that, under location shift, the relative difference in time between two landmark time points, such as the peak and lowest point in the infection density or last week compared to this week, can be estimated from the data.

## RESULTS

The National Health Commission of the People’s Republic of China has published daily numbers of confirmed cases of COVID-19 since January 20, 2020; see http://www.nhc.gov.cn/xcs/yqtb/list_gzbd.shtml for all news releases. We analyze data from areas outside the Hubei Province during the 8-week period between January 20 and March 15. Note that March 15 was selected because the majority of the newly confirmed cases have been imported cases afterwards. The daily numbers of confirmed cases are graphically depicted in Figure 2A. It can be observed that the daily number of confirmed cases reached its peak on February 15 with 890 new cases, 11 days after the lockdown of Wuhan City on January 23. The spike of 261 new cases on February 20 was due to delayed reports of outbreaks in two prisons.

**Figure 2:**
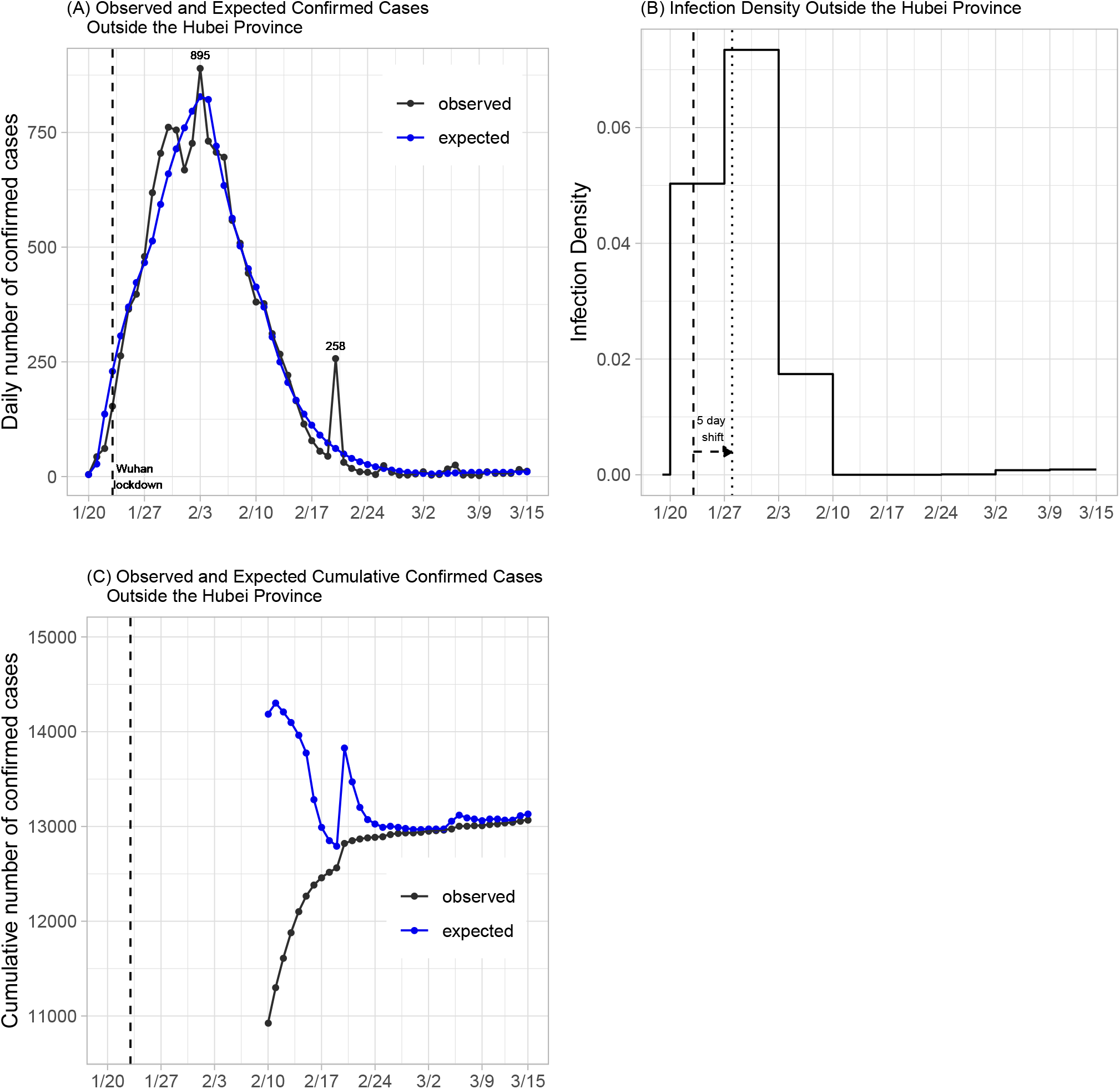
Plots of daily number of confirmed cases and infection density of confirmed cases in areas outside of the Hubei Province. Dashed and dotted lines indicate, respectively, the landmark time of Wuhan lockdown before and after accounting for a 5-day location shift.

We model the time from infection to diagnosis using the Weibull distribution, which has been shown a reasonable approximation for the incubation time in infectious disease research. As argued before, the stationarity assumption for the time to diagnosis, that is, the distribution of duration between infection to diagnosis does not depend on the time of infection, should hold approximately in cases diagnosed outside the Hubei Province. Moreover, for modeling the intensity function *ϕ*(*t*, ***α***), we assume a step function with jump discontinuities every 7 days starting from January 20 (day 0), so that the risk of infection is constant within each week. Since the first two cases diagnosed outside of Hubei were reported to have visited Wuhan on January 7 and 9 and developed symptoms on January 13 and 14, respectively, we set *τ*_0_ to January 1, 2020 to account for the possible infection period. Specifically, the incidence density function among cases infected before March 15 is of the form

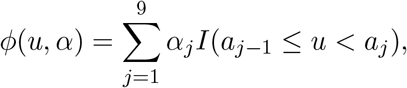

where *a*_0_ = −19, *a*_*j*_ = 7 ×(*j −*1), *j* = 1, …, 9. Note that we require 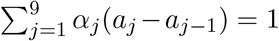 to ensure that *ϕ*(*u, α*) is a proper density function.

The back-calculation method estimates a total of 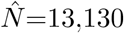 individuals (standard error [SE] 15; 95% confidence interval [CI] 13,101–13,159) who were infected before March 15. This includes 13,071 confirmed cases on and before March 15, which means that we expect 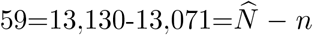 additional cases to be confirmed after March 15, should there be no new infections. The expected numbers of new cases under the fitted model are shown in Figure 2A. The predicted numbers track well with the observed data, suggesting that the proposed model fits well. The maximum likelihood estimates of the shape and size parameters for the Weibull distribution are 1.03 (SE 0.004) and 5.39 (SE 0.17), respectively. This corresponds to a median of 3.8 days (SE 0.12). As discussed before, the distribution of the time to diagnosis is estimated subject to a location shift. Recently, [9] analyzed the length-biased incubation time from 1211 confirmed COVID-19 cases who left Wuhan before the lockdown and reported a median incubation period of 8.13 days (95% CI 7.37–8.91). This suggests a location shift factor of about 5 days in our estimates.

What’s also of interest is the estimate of incidence density after the lockdown of Wuhan. The maximum likelihood estimates for *a*_*j*_’s in the 9 selected time intervals, including the time interval preceding January 20, are (2.56 × 10^*−*5^, 5.03 × 10^*−*2^, 7.34 × 10^*−*2^, 1.74 × 10^*−*2^, 7.72 × 10^*−*12^, 3.55 × 10^*−*14^, 6.75 × 10^*−*5^, 7.92 × 10^*−*4^, 9.05 × 10^*−*4^) with standard errors (1.08×10^*−*5^, 1.53×10^*−*3^, 1.29×10^*−*3^, 1.40×10^*−*3^, 4.93×10^*−*4^, 1.07×10^*−*11^, 1.21×10^*−*4^, 1.77× 10^*−*4^, 2.64 ×10^*−*4^). By definition, these estimates are for infected individuals who have been or will be diagnosed. Since the number of diagnosed individuals is proportional to the total number of infected (including those who were asymptomatic), the values provide estimates of relative risk among all infected individuals. After accounting for a location shift factor of 5 days, the peak of infection occurred immediately after the lockdown of Wuhan city (Figure 2B). Moreover, the incidence increases 46% during the week of Wuhan lockdown from the preceding week, but dropped 76% in the next week, suggesting that the travel ban that started from Wuhan and its neighboring cities had initially increased the disease spread in other provinces, but was able to effectively slow down the spread of the disease in just two weeks.

Finally, continuous assessment of potential new cases to be diagnosed in the near future can be implemented by applying the back-calculation algorithm using up-to-date data. Figure 2C shows the projected number of confirmed cases with data available up to that time point, assuming no new infections afterward. The difference between the expected and observed cumulative number of confirmed cases gives the near-term prediction of additional cases to be diagnosed. Since the prediction is relatively unstable for the first few weeks of the epidemic due to the limited amount of data, we only perform prediction after February 10, that is, the fourth week into the epidemic. As shown in Figure 2C, the prediction obtained after February 24 is very close to the total number of confirmed cases at the end of the study period (March 15). Ignoring the spike occurred around February 20, which was most likely caused by the delayed report of cases in two prisons, the prediction algorithm performs reasonably well after February 17. This shows that the back-calculation algorithm can potentially provide a useful utility for the planning of health care allocation, especially for an epidemic that is still growing.

## DISCUSSION

Different countries have taken different measures in response to the novel coronavirus, and there has been a continuing heated debate on whether aggressive COVID-19 control measures cost more than they are worth. Among all countries, China has imposed the most sweeping restrictions in response to COVID-19. The authorities locked down cities, restricted movements of millions and suspended business operations to prevent further outbreak of the disease. South Korea, another country on the front-line of the epidemic, adopted the “test and trace” strategy to aggressively test people for the disease and quarantine those who tested positive, so that the rest of the population can go about their daily lives. Both countries have observed a significant decline in the number of confirmed cases. However, to the best of our knowledge, no formal quantitative evaluation of the effect of these aggressive control measures on the incidence of infection has been conducted. This research aims to provide some preliminary evidence on the effectiveness of these measures by analyzing data from China outside of the Hubei province. We conclude that the extreme measures undertaken by the Chinese government have effectively slowed down the spread of the disease outside of the Hubei province in about two weeks. We also demonstrate that the back-calculation algorithm can be used to estimate the number of infected individuals to be diagnosed in the near future. This provides a useful utility to guide the planning of medical resource allocation in the middle of the epidemic.

## Data Availability

The National Health Commission of the People's Republic of China has published daily numbers of confirmed cases of COVID-19 since January 20, 2020; see http://www.nhc.gov.cn/xcs/yqtb/list_gzbd.shtml for all news releases.

